# Rapid disappearance of influenza following the implementation of COVID-19 mitigation measures in Hamilton, Ontario

**DOI:** 10.1101/2020.11.27.20240036

**Authors:** Kevin Zhang, Avika Misra, Patrick J. Kim, Seyed M. Moghadas, Joanne M. Langley, Marek Smieja

## Abstract

**Background:** Public health measures, such as social distancing and closure of schools and non-essential services, were rapidly implemented in Canada to interrupt the spread of the novel coronavirus disease 2019 (COVID-19).

**Objective:** We sought to investigate the impact of mitigation measures during the spring wave of COVID-19 on the incidence of other laboratory-confirmed respiratory viruses in Hamilton, Ontario.

**Methods:** All nasopharyngeal swab specimens (n = 57,503) submitted for routine respiratory virus testing at a regional laboratory serving all acute-care hospitals in Hamilton, Ontario between January 2010 and June 2020 were reviewed. Testing for influenza A/B, respiratory syncytial virus, human metapneumovirus, parainfluenza I–III, adenovirus and rhinovirus/enterovirus was done routinely using a laboratory-developed polymerase chain reaction multiplex respiratory viral panel. A Bayesian linear regression model was used to determine the trend of positivity rates of all influenza samples for the first 26 weeks of each year from 2010 to 2019. The mean positivity rate of Bayesian inference was compared with the weekly reported positivity rate of influenza samples in 2020.

**Results:** The positivity rate of influenza in 2020 diminished sharply following the population-wide implementation of COVID-19 interventions. Weeks 12-26 reported 0% positivity for influenza, with the exception of 0.1% reported in week 13.

**Conclusions:** Public health measures implemented during the COVID-19 pandemic were associated with a reduced incidence of other respiratory viruses and should be considered to mitigate severe seasonal influenza and other respiratory virus pandemics.

## Introduction

The novel coronavirus disease 2019 (COVID-19) has led to devastating global morbidity and mortality (1). Restrictive public health measures have helped to mitigate COVID-19 transmission (2,3), but have led to widespread disruptions to the economy (4,5), trade (6), and education (7). Following the declaration of COVID-19 as a pandemic on March 11 by the World Health Organization (8), the province of Ontario, Canada announced the closure of all schools and non-essential workplaces (9,10). Months later, public health measures, such as social distancing and mask-wearing, continue to be in place to reduce the toll associated with the COVID-19 pandemic (11).

Public health measures have reduced the transmission of severe acute respiratory syndrome coronavirus 2 (SARS-CoV-2) in Ontario (3). In some jurisdictions, these measures have also been associated with a lower incidence of other respiratory virus infections (12,13). We performed a time-series analysis, using a hierarchical regression model, to determine the timelines and positivity rates of influenza A and B viruses from 2010 to 2019 in an urban center in Ontario, and compare them to those of 2020 prior to and following the implementation of COVID-19 interventions in response to initial outbreaks.

## Methods

### Sampling and testing

We reviewed all nasopharyngeal swab specimens (n = 57,503) submitted for routine respiratory virus testing at a regional laboratory serving all acute-care hospitals in Hamilton, Ontario between January 2010 and June 2020.

Testing was done using a Taqman real-time reverse transcription polymerase chain reaction multiplex respiratory viral panel, developed by the Hamilton Regional Laboratory Medicine Program, for influenza A/B, respiratory syncytial virus, human metapneumovirus, parainfluenza I–III, adenovirus, and rhinovirus/enterovirus. On March 16, 2020, parainfluenza II was replaced by the SARS-CoV-2 virus. Sample RNA extraction and amplification were primarily performed on the bioMérieux NucliSENS easyMag and QIAGEN Rotor-Gene Q, respectively, from 2010-2019 and primarily performed on the BD MAX System from July 2019-2020. Clinical results were validated by experienced staff and recorded into a laboratory information system, following standard operating procedures.

### Data

A respiratory virus database with all test results and demographic information is updated weekly, and has been in place since 2010. A 10-years datacut with basic demographic information (age, sex, postal code, date, facility, accession number) and test results were exported from the laboratory database on June 29, 2020. The database included only samples sent for multiplex testing. Laboratory test results were filtered by postal code to exclude samples from persons living outside of Hamilton, Ontario. Figure 1 shows the positivity rates of influenza A/B in the database for different age groups.

**Figure 1.**
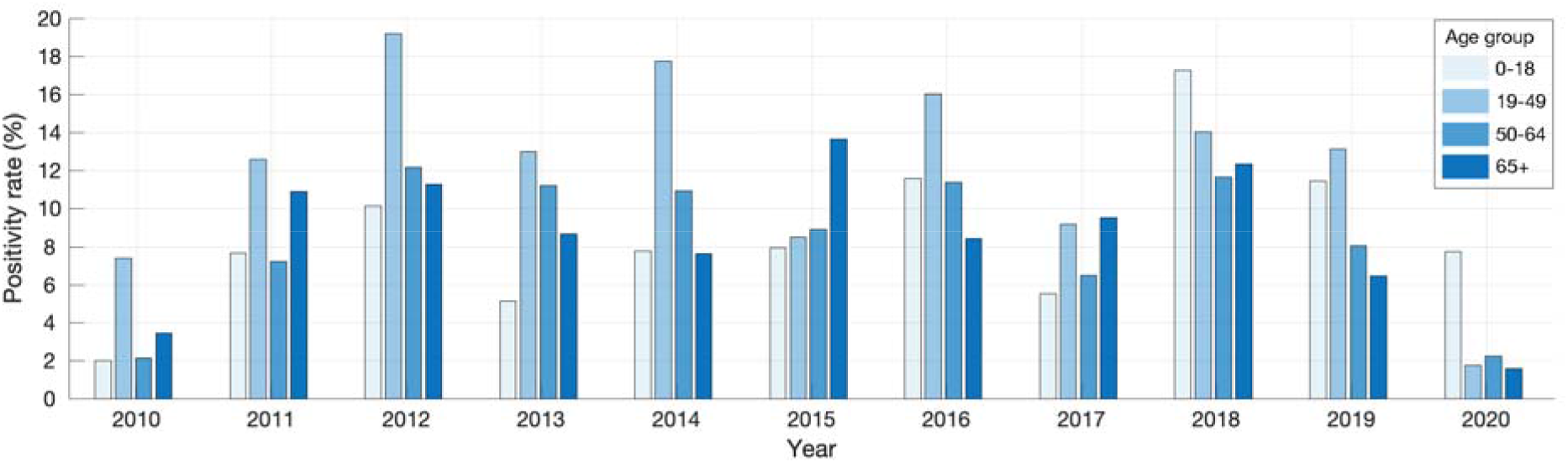
Positivity rates of influenza A/B in 2010-2020 for different age groups in Hamilton, Ontario (0 to 18 years, 19 to 49 years, 50 to 64 years and 65 years of age and older).

### Ethics approval

The study was approved by the Hamilton Integrated Research Ethics Board (Project: 07-2923). The study was categorized as minimal risk, defined as no potential for negative impact on the health and safety of the participant, and waiver of individual consent for participation was obtained.

### Statistical analysis

We used a Bayesian linear regression model with uninformative prior distributions to determine the trend of positivity rates of all influenza A/B samples for the first 26 weeks of each year from 2010-2019 (Appendix: Table A1). We then compared the mean positivity rate of Bayesian inference with the weekly reported positivity rate of influenza samples in 2020 (Appendix: Table A2).

The hierarchical regression model has the form

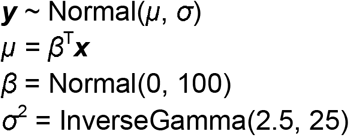

where ***y*** represents the positivity rate over the first 26 weeks (variable ***x***) of each year from 2010-2019. All parameters were sampled using Markov Chain Monte Carlo (MCMC) simulations in three independent chains. Each chain consisted of 10,000 iterations, with a burn-in period of 1,000 iterations and a thinning factor of 5. To assess convergence, we inspected the trace plots and applied the Gelman-Rubin convergence test by computing the potential scale reduction factors (PSRF). All PSRF values were computed to be less than 1.1 (and remained close to 1), indicating the convergence of the model parameters to their posterior distributions. We used the posterior distributions of the parameters (β_1_, β_2_, σ) from our Bayesian analysis to derive mean estimates and credible intervals (Appendix: Table A3) by employing the method of Highest Posterior Density (14).

## Results

A description of individuals included in our study is provided in Table 1. A total of 48,459 patients were tested for respiratory viruses in Hamilton, Ontario in 2010-2019, of which 49.3% (n = 23,898) were male and 30.6% (n = 14,818) were children under 18 years of age. The bimodal age distribution had a median age of adults of 72.4 years (IQR: 59.4 - 83.5) and 1.5 years among children (IQR: 0.4 - 4.4). A median of 4,626 (IQR: 3,376 - 5,936) samples were tested each year, with a mean influenza positivity rate of 9.6% (SD: 2.9%). Mean percent positivity was also calculated for respiratory syncytial virus (6.9%, SD: 1.5%), metapneumovirus (2.8%, SD: 0.4%), parainfluenza (3.2%, SD: 0.6%), adenovirus (1.0%, SD: 0.6%), and rhinovirus/enterovirus (8.0%, SD: 5.5%). 9,044 patients were tested for respiratory viruses in 2020, of which 2.5% were positive for influenza. The percent positivity of other respiratory viruses ranged from 0.1% (parainfluenza) to 0.9% (respiratory syncytial virus and rhinovirus/enterovirus).

**Table 1.** Demographics, sample size, and positivity rate of laboratory-confirmed respiratory viruses in Hamilton, Ontario in 2010-2019 (n = 48,459) and 2020 (n = 9,044).

|  | Median (IQR) |  |
| --- | --- | --- |
| Demographics | 2010-2019 | 2020 |
| Age in years, adults | 72.4 (59.4 - 83.5) | 63.0 (46.1 - 77.2) |
| Age in years, children | 1.5 (0.4 - 4.4) | 1.9 (0.5 - 6.0) |
| Male | 23,898 (49.3%) | 4,073 (45.0%) |
| Adults | 33,641 (69.4%) | 7,983 (88.3%) |
| Children | 14,818 (30.6%) | 1,061 (11.7%) |
| Respiratory virus samples | Median (IQR) |  |
| Samples per year | 4,626 (3,376 - 5,936) | 9,044 |
| Positivity rate | Mean (SD) |  |
| Influenza | 9.6% (2.9%) | 2.5% |
| Respiratory syncytial virus | 6.9% (1.5%) | 0.9% |
| Metapneumovirus | 2.8% (0.4%) | 0.4% |
| Parainfluenza | 3.2% (0.6%) | 0.1% |
| Adenovirus | 1.0% (0.6%) | 0.2% |
| Rhinovirus/enterovirus | 8.0% (5.5%) | 0.9% |

Figure 2 illustrates the mean positivity rate derived from posterior distributions of parameters in the Bayesian linear regression model using positivity rates reported for 2010-2019 (black curve). The positivity rate of influenza in 2020 (red curve) was highest at 17.7% in week 1, and dropped below the 95% CrI for the preceding 10 years after the first week, with an ensuing declining trend (Figure 2; Appendix: Table A2). Following the implementation of COVID-19 interventions during week 12 (from March 12, 2020; grey bar in Figure 2), the positivity rate of influenza diminished sharply and remained at 0% for weeks 12-26, with the exception of 0.1% reported in week 13.

**Figure 2.**
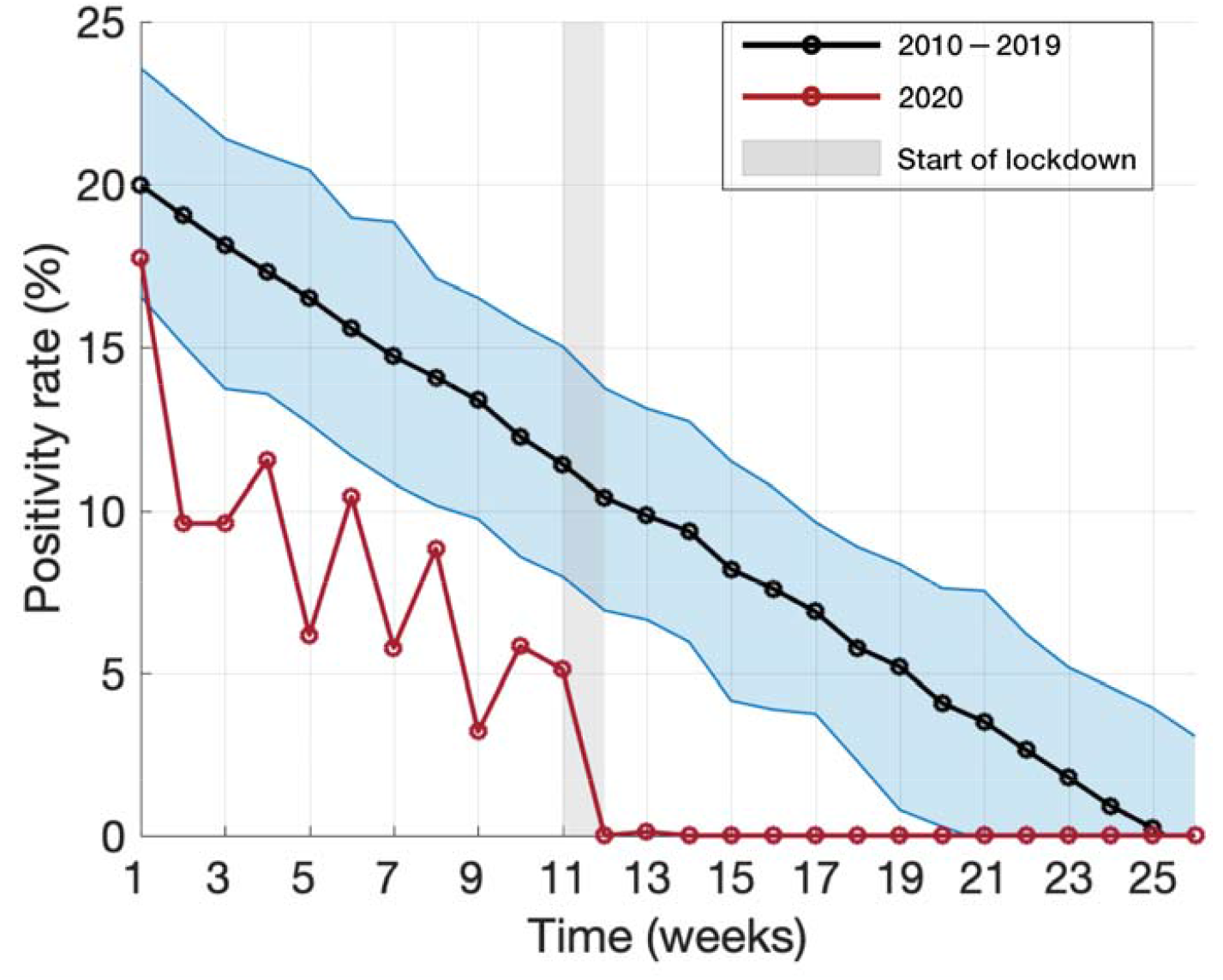
Bayesian inference for the mean positivity rate (black curve) and its 95% CrI (blue shaded area) of influenza A and B for the first 26 weeks in 2010-2019. The red curve shows the positivity rate of influenza A and B for 2020, with the shaded grey bar indicating the start of COVID-19 lockdown.

## Discussion

Public health measures have been used to interrupt spread of influenza during pandemics, with effect. For example, school closures and social distancing during the 2009 H1N1 pandemic in Mexico resulted in a 27-29% reduction in influenza transmission during the spring wave (15). During the 1957-1958 influenza pandemic, school closures contributed to reducing the attack rate by over 90% (16). Similarly, following the implementation of COVID-19 mitigation measures, the influenza positivity rate was suppressed in the United States (12,13). Our results suggest that COVID-19 public health measures may have contributed to a substantial disruption of the spread of influenza in Hamilton, Ontario.

The 2020 influenza season was observed to be relatively mild in Hamilton, Ontario, as compared to previous seasons (Appendix: Table A1, A2). However, the lower positivity rate observed in our analysis (Figure 2), may be attributed to several factors including voluntary precautions taken by individuals as a result of initial news reporting of the spread of COVID-19 in China and internationally, normal seasonal variation, or due to changes in sampling behaviour and diagnostic testing. For the 2010-2019 winter influenza season, the median influenza positivity rate reached 0% by week 23. In 2020, however, after the implementation of COVID-19 mitigation measures, percent positivity for influenza dropped precipitously to 0% in week 12. The Centers for Disease Control and Prevention reported similar findings through their weekly influenza surveillance system, in which the percent positivity for influenza decreased from 7.5% in week 12 to 1.0% in week 14. This abrupt change, without another explanation, suggests that COVID-19 mitigation measures may have reduced the spread of laboratory-confirmed influenza in the United States (12,13). Moreover, the positivity rates for respiratory syncytial virus, metapneumovirus, parainfluenza, adenovirus, and rhinovirus/enterovirus were reported to be 0% by week 14 of 2020 (Appendix: Table A2), suggesting that public health measures could have also suppressed the transmission of other respiratory viruses.

Understanding the effect of COVID-19 interventions on other communicable diseases requires further study. However, a number of explicators may be considered to describe the rapid interruption in transmission chains of influenza compared to COVID-19 with the pressure exerted by public health measures. First, there is relatively strong cross-immunity for influenza virus strains during seasonal epidemics, in addition to population immunity conferred by vaccination (17,18). In contrast, the population was naive to SARS-CoV-2, and still remains largely susceptible in the absence of vaccination. Furthermore, there are also major differences in the epidemiological characteristics between influenza and COVID-19 that influence the outcomes of interventions (19). For example, the transmissibility of influenza has been estimated in the range 1.2-1.8 (20), which is lower than the initial estimates greater than 2 for COVID-19 in most settings (21,22). The average incubation period of 5.2 days for COVID-19 (21) is significantly longer than the same period for influenza A, which is estimated to be 1.4 days (23). Moreover, the pre-symptomatic period is longer and more infectious in COVID-19 than in influenza (24,25). Future studies will need to account for these factors when evaluating the effect of interventions against emerging infectious diseases.

The findings of our study should be interpreted in the context of study limitations. First, respiratory samples were not collected systematically, but rather they were obtained as part of routine clinical care. As such, the samples may not fully represent the prevalence of respiratory viruses in the region. It is also possible that clinicians may not have strictly followed hospital infection control policy and failed to sample patients who otherwise would have been eligible. Furthermore, sampling behaviour may have changed during the early stage of COVID-19 spread in Canada. However, these factors are unlikely to change our conclusions due to the near-elimination of the absolute number of laboratory-confirmed respiratory virus cases, despite the large increase in testing which accompanied concern for COVID-19 in the community.

Our findings suggest that efforts to control the COVID-19 pandemic may have had additional benefits in suppressing the transmission of other respiratory viruses in Hamilton, Ontario. Mitigation strategies, such as social distancing, mask-wearing, and school closures could play an important role in combating future seasonal respiratory viruses and emerging infectious diseases with pandemic potential.

## Data Availability

The datasets generated for the study are not publicly available due to the presence of personally identifiable information. Aggregate data, however, are presented in the manuscript within the Table, Figure, and Supplementary File and are available from the corresponding author on reasonable request.

## Authors’ statement

KZ, AM, PJK, SMM, and MS contributed to the conception and design of the work. KZ, SMM, JML, and MS contributed to the acquisition of data, analysis, and interpretation of results. All authors drafted, read, and approved the final manuscript.

## References

1. Johns Hopkins University. COVID-19 Map [Internet]. Johns Hopkins Coronavirus Resource Center. [cited 2020 Sep 1]. Available from: https://coronavirus.jhu.edu/map.html

2. Zhang K, Vilches TN, Tariq M, Galvani AP, Moghadas SM. The impact of mask-wearing and shelter-in-place on COVID-19 outbreaks in the United States. International Journal of Infectious Diseases [Internet]. 2020 Dec [cited 2020 Dec 23];101:334–41. Available from: https://www.ijidonline.com/article/S1201-9712(20)32204-9/fulltext

3. Public Health Ontario. COVID-19 Seroprevalence in Ontario: March 27, 2020 to June 30, 2020. 2020;10.

4. Fernandes N. Economic Effects of Coronavirus Outbreak (COVID-19) on the World Economy [Internet]. Rochester, NY: Social Science Research Network; 2020 Mar [cited 2020 Aug 17]. Report No.: ID 3557504. Available from: https://papers.ssrn.com/abstract=3557504

5. Jones L, Palumbo D, Brown D. Coronavirus: A visual guide to the economic impact. BBC News [Internet]. 2020 Jun 30 [cited 2020 Aug 17]; Available from: https://www.bbc.com/news/business-51706225

6. World Trade Organization. COVID-19 and world trade [Internet]. [cited 2020 Aug 17]. Available from: https://www.wto.org/english/tratop_e/covid19_e/covid19_e.htm

7. Dorn E, Hancock B, Sarakatsannis J, Viruleg E. Achievement gap and coronavirus | McKinsey [Internet]. [cited 2020 Aug 17]. Available from: https://www.mckinsey.com/industries/public-and-social-sector/our-insights/covid-19-and-student-learning-in-the-united-states-the-hurt-could-last-a-lifetime

8. World Health Organization. WHO Director-General’s opening remarks at the media briefing on COVID-19 - 11 March 2020 [Internet]. [cited 2020 Aug 17]. Available from: https://www.who.int/dg/speeches/detail/who-director-general-s-opening-remarks-at-the-media-briefing-on-covid-1911-march-2020

9. Rodrigues G. Ontario government declares state of emergency amid coronavirus pandemic [Internet]. Global News. [cited 2020 Aug 17]. Available from: https://globalnews.ca/news/6688074/ontario-doug-ford-coronavirus-covid-19-march-17/

10. Vogel L. COVID-19: A timeline of Canada’s first-wave response | CMAJ News [Internet]. [cited 2020 Aug 17]. Available from: https://cmajnews.com/2020/06/12/coronavirus-1095847/

11. Government of Ontario. Reopening Ontario | Ontario.ca [Internet]. [cited 2020 Aug 17]. Available from: https://www.ontario.ca/page/reopening-ontario

12. Centers for Disease Control and Prevention. INFLUENZA Isolates Data Table, Season 2019-2020 [Internet]. [cited 2020 Aug 17]. Available from: https://www.cdc.gov/flu/weekly/weeklyarchives2019-2020/data/whoAllregt_cl32.html

13. Blum K. Was Social Distancing a Help in Slowing the Flu Season? [Internet]. [cited 2020 Aug 17]. Available from: https://www.idse.net/Covid-19/Article/04-20/Was-Social-Distancing-a-Help-in-Slowing-the-Flu-Season-/57839?ses=ogst

14. Chen M-H, Shao Q-M. Monte Carlo Estimation of Bayesian Credible and HPD Intervals. Journal of Computational and Graphical Statistics [Internet]. 1999 Mar [cited 2020 Oct 6];8(1):69–92. Available from: http://www.tandfonline.com/doi/abs/10.1080/10618600.1999.10474802

15. Stephenson J. Social Distancing Helpful in Mexico During Flu Pandemic. JAMA [Internet]. 2011 Jun 22 [cited 2020 Aug 17];305(24):2509–2509. Available from: https://jamanetwork.com/journals/jama/fullarticle/646743

16. Glass RJ, Glass LM, Beyeler WE, Min HJ. Targeted Social Distancing Designs for Pandemic Influenza. Emerg Infect Dis [Internet]. 2006 Nov [cited 2020 Aug 17];12(11):1671–81. Available from: https://www.ncbi.nlm.nih.gov/pmc/articles/PMC3372334/

17. Valkenburg SA, Rutigliano JA, Ellebedy AH, Doherty PC, Thomas PG, Kedzierska K. Immunity to seasonal and pandemic influenza A viruses. Microbes and Infection [Internet]. 2011 May [cited 2020 Dec 24];13(5):489–501. Available from: https://linkinghub.elsevier.com/retrieve/pii/S1286457911000372

18. Krammer F. The human antibody response to influenza A virus infection and vaccination. Nat Rev Immunol [Internet]. 2019 Jun [cited 2020 Dec 24];19(6):383–97. Available from: http://www.nature.com/articles/s41577-019-0143-6

19. Abdollahi E, Haworth-Brockman M, Keynan Y, Langley JM, Moghadas SM. Simulating the effect of school closure during COVID-19 outbreaks in Ontario, Canada. BMC Med [Internet]. 2020 Dec [cited 2020 Dec 24];18(1):230. Available from:https://bmcmedicine.biomedcentral.com/articles/10.1186/s12916-020-01705-8

20. Biggerstaff M, Cauchemez S, Reed C, Gambhir M, Finelli L. Estimates of the reproduction number for seasonal, pandemic, and zoonotic influenza: a systematic review of the literature. BMC Infect Dis [Internet]. 2014 Dec [cited 2020 Dec 24];14(1):480. Available from: https://bmcinfectdis.biomedcentral.com/articles/10.1186/1471-2334-14-480

21. Li Q, Guan X, Wu P, Wang X, Zhou L, Tong Y, et al. Early Transmission Dynamics in Wuhan, China, of Novel Coronavirus–Infected Pneumonia. N Engl J Med [Internet]. 2020 Mar 26 [cited 2020 Dec 24];382(13):1199–207. Available from: http://www.nejm.org/doi/10.1056/NEJMoa2001316

22. Wu JT, Leung K, Leung GM. Nowcasting and forecasting the potential domestic and international spread of the 2019-nCoV outbreak originating in Wuhan, China: a modelling study. The Lancet [Internet]. 2020 Feb [cited 2020 Dec 24];395(10225):689–97. Available from: https://linkinghub.elsevier.com/retrieve/pii/S0140673620302609

23. Lessler J, Reich NG, Brookmeyer R, Perl TM, Nelson KE, Cummings DA. Incubation periods of acute respiratory viral infections: a systematic review. The Lancet Infectious Diseases [Internet]. 2009 May [cited 2020 Dec 24];9(5):291–300. Available from: https://linkinghub.elsevier.com/retrieve/pii/S1473309909700696

24. He X, Lau EHY, Wu P, Deng X, Wang J, Hao X, et al. Temporal dynamics in viral shedding and transmissibility of COVID-19. Nature Medicine [Internet]. 2020 May [cited 2020 Oct 27];26(5):672–5. Available from: https://www.nature.com/articles/s41591-020-0869-5

25. Moghadas SM, Fitzpatrick MC, Sah P, Pandey A, Shoukat A, Singer BH, et al. The implications of silent transmission for the control of COVID-19 outbreaks. PNAS [Internet]. 2020 Jul 28 [cited 2020 Oct 27];117(30):17513–5. Available from: https://www.pnas.org/content/117/30/17513

