## Supplementary File for "Rapid disappearance of influenza following the implementation of COVID-19 mitigation measures in Hamilton, Ontario"


**Table A1.** Percent positivity of laboratory-confirmed influenza in Hamilton, Ontario in weeks 1-26 of 2010-2019.

| Week | Mean | Median | Quartile 1 | Quartile 3 | Standard Deviation |
|---|---|---|---|---|---|
| 1 | 18.5 | 21.1 | 11.0 | 26.2 | 10.3 |
| 2 | 16.0 | 15.6 | 3.6 | 29.8 | 11.2 |
| 3 | 17.4 | 21.0 | 5.4 | 26.1 | 10.0 |
| 4 | 16.5 | 12.2 | 5.8 | 26.6 | 11.9 |
| 5 | 16.8 | 17.3 | 8.8 | 24.9 | 10.3 |
| 6 | 17.5 | 16.7 | 12.7 | 23.8 | 9.2 |
| 7 | 15.2 | 16.0 | 7.4 | 21.5 | 8.8 |
| 8 | 15.9 | 15.2 | 6.8 | 23.8 | 9.3 |
| 9 | 14.6 | 13.9 | 8.2 | 20.7 | 9.1 |
| 10 | 14.8 | 14.2 | 5.8 | 20.0 | 10.1 |
| 11 | 12.7 | 11.2 | 3.9 | 21.6 | 8.5 |

| | | | | | |
|---|---|---|---|---|---|
| 12 | 10.2 | 10.9 | 5.1 | 16.0 | 6.2 |
| 13 | 11.4 | 10.9 | 5.6 | 18.4 | 6.8 |
| 14 | 11.5 | 9.8 | 3.6 | 20.3 | 8.4 |
| 15 | 8.5 | 7.6 | 4.1 | 11.7 | 5.8 |
| 16 | 7.4 | 5.4 | 2.5 | 13.1 | 5.9 |
| 17 | 7.3 | 5.8 | 2.2 | 11.9 | 5.6 |
| 18 | 5.5 | 6.3 | 2.2 | 7.1 | 3.4 |
| 19 | 3.2 | 2.0 | 0.9 | 5.8 | 3.2 |
| 20 | 3.3 | 1.6 | 0.7 | 6.3 | 3.3 |
| 21 | 1.9 | 0.7 | 0.0 | 2.4 | 3.0 |
| 22 | 1.4 | 0.7 | 0.0 | 3.0 | 1.5 |
| 23 | 1.2 | 0.0 | 0.0 | 2.1 | 1.9 |
| 24 | 0.8 | 0.0 | 0.0 | 1.9 | 1.1 |
| 25 | 0.3 | 0.0 | 0.0 | 0.3 | 0.6 |
| 26 | 0.0 | 0.0 | 0.0 | 0.0 | 0.0 |

**Table A2.** Percent positivity of laboratory-confirmed influenza, respiratory syncytial virus, metapneumovirus, parainfluenza, adenovirus, and rhinovirus/enterovirus in Hamilton, Ontario in weeks 1-26 of 2020.

| Week | Influenza Percent Positivity | Respiratory Syncytial Virus Percent Positivity | Metapneumovirus Percent Positivity | Parainfluenza Percent Positivity | Adenovirus Percent Positivity | Rhinovirus/ Enterovirus Percent Positivity |
|---|---|---|---|---|---|---|
| 1 | 17.7 | 5.9 | 0.5 | 0.0 | 0.5 | 2.2 |
| 2 | 9.6 | 5.6 | 0.8 | 0.4 | 0.8 | 1.6 |
| 3 | 9.6 | 1.8 | 0.9 | 0.4 | 0.4 | 3.9 |
| 4 | 11.5 | 5.8 | 0.8 | 0.0 | 0.4 | 1.2 |
| 5 | 6.2 | 4.5 | 0.7 | 0.0 | 0.3 | 2.4 |
| 6 | 10.4 | 2.3 | 1.2 | 0.4 | 0.8 | 1.5 |
| 7 | 5.8 | 1.9 | 1.0 | 0.0 | 1.0 | 1.4 |
| 8 | 8.8 | 2.4 | 2.4 | 0.0 | 0.0 | 2.4 |
| 9 | 3.2 | 0.5 | 0.5 | 0.0 | 0.5 | 0.0 |
| 10 | 5.8 | 1.5 | 1.9 | 1.0 | 1.0 | 4.4 |

| | | | | | | |
|---|---|---|---|---|---|---|
| 11 | 5.1 | 0.6 | 2.4 | 0.4 | 0.4 | 6.1 |
| 12 | 0.0 | 0.0 | 0.5 | 0.2 | 0.0 | 0.2 |
| 13 | 0.1 | 0.0 | 0.1 | 0.1 | 0.0 | 0.2 |
| 14 | 0.0 | 0.0 | 0.0 | 0.0 | 0.0 | 0.0 |
| 15 | 0.0 | 0.0 | 0.0 | 0.0 | 0.0 | 0.0 |
| 16 | 0.0 | 0.0 | 0.0 | 0.0 | 0.0 | 0.0 |
| 17 | 0.0 | 0.0 | 0.0 | 0.0 | 0.0 | 0.0 |
| 18 | 0.0 | 0.0 | 0.0 | 0.0 | 0.0 | 0.0 |
| 19 | 0.0 | 0.0 | 0.0 | 0.0 | 0.0 | 0.0 |
| 20 | 0.0 | 0.0 | 0.0 | 0.0 | 0.0 | 0.0 |
| 21 | 0.0 | 0.0 | 0.0 | 0.0 | 0.0 | 0.0 |
| 22 | 0.0 | 0.0 | 0.0 | 0.0 | 0.0 | 0.0 |
| 23 | 0.0 | 0.0 | 0.0 | 0.0 | 0.0 | 0.0 |
| 24 | 0.0 | 0.0 | 0.0 | 0.0 | 0.0 | 0.0 |
| 25 | 0.0 | 0.0 | 0.0 | 0.0 | 0.0 | 0.0 |
| 26 | 0.0 | 0.0 | 0.0 | 0.0 | 0.0 | 0.0 |

**Table A3.** Estimated model parameters from Bayesian inference.

| Parameter | Mean | 95% CrI |
|---|---|---|
| $\beta_1$ | 13.494 | -0.891, 21.695 |
| $\beta_2$ | 0.114 | -0.888, 1.985 |
| $\sigma$ | 4.005 | 1.932, 5.102 |
